# Molecular pathways in post-colonoscopy versus detected colorectal cancers: results from a nested case-control study

**DOI:** 10.1101/2021.06.28.21259642

**Authors:** Roel M.M. Bogie, Chantal M.C. le Clercq, Quirinus J.M. Voorham, Martijn Cordes, Daoud Sie, Christian Rausch, Evert van den Broek, Sara D.J. de Vries, Nicole C.T. van Grieken, Robert G. Riedl, Prapto Sastrowijoto, Ernst-Jan Speel, Rein Vos, Bjorn Winkens, Manon van Engeland, Bauke Ylstra, Gerrit A. Meijer, Ad A.M. Masclee, Beatriz Carvalho

## Abstract

**Background:** Post-colonoscopy colorectal cancers (PCCRCs) pose a challenge in clinical practice. PCCRCs occur due to a combination of procedural and biological causes. Specific features of lesions may contribute for this. In a nested case-control study, we compared clinical and molecular features of PCCRCs and detected CRCs (DCRCs).

**Methods:** PCCRCs were defined according to the WEO 2018 classification, as cancers occurring after a complete index colonoscopy, which excluded CRC. CRCs in patients without colonoscopy or with colonoscopy >10 years before were defined as DCRCs. Whole genome chromosomal copy number changes and mutation status of genes commonly affected in CRC (including *APC, KRAS, BRAF, FBXW7, PIK3CA, NRAS, SMAD4* and *TP53*) were examined by low-coverage WGS and targeted sequencing, respectively. MSI and CIMP status were also determined.

**Results:** In total, 122 PCCRCs and 98 DCRCs with high quality DNA were examined. PCCRCs were more often located proximally in the colon (p<0.001), non-polypoid appearing (p=0.004), early stage (p=0.009), and poorly differentiated (p=0.006). PCCRCs showed similar patterns of DNA copy number changes typical of CRC, although significantly less 18q loss (FDR <0.2), compared to DCRCs. No significant differences in mutations were detected between PCCRCs and DCRCs. PCCRCs were more commonly CIMP-high (p=0.014) and MSI (p=0.029). After correction for tumour location, the only molecular difference between PCCRCs and DCRCs that remained significant was less frequent loss of 18q chromosome in PCCRCs (p=0.005).

**Conclusion:** Although PCCRCs show molecular characteristics that are common to the canonical CIN, MSI and hypermethylation pathways, molecular features associated with the sessile serrated lesions (SSLs) and non-polypoid colorectal neoplasms (CRNs) are more commonly seen in PCCRCs than in DCRCs. This and the clinical features observed in PCCRCs support the hypothesis that sessile serrated lesions and non-polypoid CRNs are contributors to the development of these cancers. In order to further reduce the occurrence of PCCRCs, the focus should be directed at improving the detection, determination and endoscopic removal of these non-polypoid CRN and SSLs.

**Clinical Trial Registration:** NTR3093 in the Dutch trial register (www.trialregister.nl)

## Background

Colonoscopy is an effective screening tool for colorectal cancer. However, in 3.7% (95% CI: 2.8-4.9%) of all colorectal carcinomas, a preceding colonoscopy did not detect the (pre-)malignant lesion.^1^ These so called post-colonoscopy colorectal cancers (PCCRCs) can be subdivided with respect to aetiology into biological factors and procedural factors.^2-4^ In previous studies, it was noted that more than half of the PCCRCs had missed lesions as most likely aetiology.^5,6^

It is hypothesized that the underlying mechanisms may differ depending on the causes of PCCRCs. Missed lesions could be the result of non-polypoid (flat) colonic lesions which are easily overlooked during endoscopy.^7^ Large flat lesions, the so-called laterally spreading tumours, frequently contain high grade dysplasia and early carcinoma.^8,9^ Resection of these lesions is more difficult, leading to higher recurrence rates.^10^ Sessile serrated lesions (SSL) are often flat and have a pale appearance, thereby increasing the risk of being missed.^11^ These lesions are thought to develop into CRC via a different molecular pathway.^12,13^ Newly developed cancers may result from a fast growing precursor lesion. Underlying molecular pathological mechanisms, such as microsatellite instability (MSI), could be involved in this more rapid development.^2,14^

Previous studies have pointed to differences in molecular profiles between PCCRCs and detected CRCs (DCRCs) with more often MSI and CpG island methylator phenotype (CIMP) in PCCRCs.^14-16^ Here, detected CRCs are defined as CRCs found in patients without previous colonoscopy or with colonoscopy >10 years ago. Several studies showed that after correcting for tumour location, no differences were found in genetics between PCCRCs and DCRCs.^17,18^ In the current study, next to MSI and methylation status, whole genome DNA copy number changes and mutations in CRC-related genes was performed, in order to assess the biological pathways involved in PCCRCs. Based on the WEO classification for PCCRCs, we compared PCCRCs to DCRCs, in a nested case-control study. Second, we compared the subgroup of PCCRCs with probable biological aetiology with detected CRCs, so that procedural causes would not confound the biology behind PCCRCs. We hypothesize that PCCRCs have a molecular profile that is different from DCRCs, presumably more similar to non-polypoid and/or sessile serrated precursor lesions.

## Material and methods

### Study population

All colorectal cancers detected between January 1^st^ of 2001 and December 31^st^ of 2010 were collected in three large-volume hospitals (one university and two large general teaching hospitals) in the region of South Limburg, the Netherlands.^5^ An electronic pathology database was used to identify all CRCs and this was crosschecked with the Dutch Cancer Registration. Patients with hereditary CRC, inflammatory bowel disease (IBD) or a history of previous CRC were excluded. For each case, data of the last colonoscopy were retrieved from patient files in the three local hospitals. Based on its geography, the South Limburg region is frequently used for population-based studies. It is characterised by a stable population over time, as shown by a low net migration rate (0.8 per 1000 inhabitants per year). ^19^ The study was approved by the Medical Ethical Committee of the Maastricht University Medical Centre, which waived the need for informed consent because of the retrospective character and absence of possible consequences for individual diagnosis. The study is registered as study NTR3093 in the Dutch trial register (www.trialregister.nl).

### Definitions

The WEO consensus statement was published in 2018, after the period of data collection from 2001-2010.^20^ Since variables as caecal intubation, faecal contamination and whether a CRC was detected in a segment with previous neoplasia were registered in the database, as well as the detailed Pabby classification,^4^ retrospective application of the WEO definition to the prospectively collected dataset was possible and has been used.

Post-colonoscopy CRCs (PCCRCs) were defined as colorectal carcinomas that were detected between 6 months and maximum of 10 years after index colonoscopy that was negative for CRC, according to the WEO guideline for PCCRC.^20^ Detected CRCs (DCRCs) were defined as CRCs without prior colonoscopy or colonoscopy >10 years before. The most likely aetiology of each PCCRC was determined. Based on this algorithm, all PCCRCs were selected from the population-based database containing all CRCs.

PCCRC cases were identified among the CRCs in the database, based on the calculated interval between diagnosis and last colonoscopy. Then, all PCCRC cases were manually checked for the most probable explanation based on the patient records. Probably missed and probably new PCCRCs were more likely related to biological factors than PCCRCs probably related to incomplete resections of lesions, no resection at all or missed lesions due to inadequate prior examination. We will further refer to those categories as biological PCCRCs and procedural PCCRCs respectively. According to WEO 2018 classification,^20^ probably missed lesions with prior adequate examination PCCRCs (<4 years after colonoscopy) can only be found after a complete (cecum visualization) colonoscopy in a well-prepared colon with no previous resection at the site of the metachronous PCCRC. These features were prospectively collected for each case.

Morphology (protruded vs flat) was based on endoscopists and pathologist’s judgement. Distal location was defined as distal from the splenic flexure. Tissues of all PCCRCs and an equal number of randomly selected DCRCs were selected for DNA analysis. To assess and (afterwards) control for the effect of all tumour features on the molecular profile a random control group, instead of a matched one (which would remove the influence of one or two features), was drawn. In addition, a matched sample would result in a smaller effective sample size in case of missing values, since the matched control (case) is then also treated as missing if the case (control) is missing.

### Material

Formalin-fixed, paraffin-embedded (FFPE) samples from the CRCs were used for DNA extraction. All data and tissues were coded. Archival material was used in compliance with the institutional ethical regulations and national guidelines.

DNA was isolated as previously described.^21^ In brief, DNA from FFPE material was isolated following macro-dissection (>70% cancer cells). A three-day incubation period with proteinase K in lysis buffer (ATL buffer, QIAmp, DNA micro-kit, Qiagen, Venlo, The Netherlands) was performed. Every day, proteinase K (10 µl of 20 ng/µl) was freshly added. DNA was isolated using the QIAmp DNA micro-kit (Qiagen) and concentrations and purity were measured on a Nanodrop ND-1000 spectrophotometer (Isogen, IJsselstein, The Netherlands).

### DNA copy number alterations analysis

DNA copy number alterations analysis was performed by low-coverage whole genome sequencing (WGS).^22^ Briefly, DNA was fragmented by sonication (Covaris S2, Woburn, MA, USA) and run on the HiSeq 2000 (Illumina, San Diego, CA, USA) on a 50 bp single-read modus using the Illumina Truseq Nano kit. DNA copy number data analysis was done as previously described.^23^

### Mutation analysis

For mutation analysis, the TruSeq Amplicon Cancer Panel (TSACP; Illumina Inc, San Diego, CA, USA) comprising 212 amplicons from 48 genes that are simultaneously amplified in a single-tube reaction, was used. Of each FFPE-DNA sample a total of 150 ng DNA (unless otherwise specified) was used as input for amplicon library preparation according to the manufacturer’s instructions. Up to 24 differently barcoded, individual sequence libraries were equimolarly pooled prior to sequencing. These multiple-sample sequence library pools were loaded either on a MiSeq Personal Sequencer (Illumina) using a MiSeq Reagent Kit v2 (300 cycles) (Illumina), according to the manufacturer’s instructions (first 28 samples), or loaded on a HiSeq2500 and run in rapid run mode, 150bp paired-end (the rest of the samples).

### MSI status analysis

MSI analysis was performed using a multiplex marker panel (MSI Multiplex System Version 1.2, Promega, Madison, WI, USA), as previously described.^24^ When two or more markers were instable, the sample was classified as microsatellite instable (MSI), all other samples were classified as microsatellite stable.

### CIMP status analysis

CpG island methylator phenotype (CIMP) in the CRC samples was determined using the CIMP panel (*CACNA1G, IGF2, NEUROG1, RUNX3* and *SOCS1*) as defined by Weisenberger et al.^25^ by nested methylation-specific PCR (MSP) using sodium bisulphite modified genomic DNA (EZ DNA methylation kit ZYMO research Co., Orange, CA, USA), as described before.^26,27^ CRCs were classified as CIMP-positive when ≥3 of the 5 CIMP markers were methylated.^25^

### Statistical analysis

Patient characteristics were analysed with descriptive statistics. To compare differences between the PCCRCs and DCRCs regarding their clinic-pathologic features, independent-samples t-test for age distribution or Chi-square test (or Fisher’s exact test, when applicable) for all categorical features were applied. P-values <0.05 were considered significant.

To analyse the genomic changes between selected groups of patients CGHtest 1.1 was used.^28^ P-values were calculated by performing a Chi-square test with 10.000 permutations. Separate analyses were run to test for gains and losses. This test procedure includes a permutation-based false discovery rate (FDR) correction for multiple testing. Alterations occurring less than 5% were a priori excluded and an FDR<0.2 was considered statistically significant.

A logistic regression model correcting for age, gender and location was applied after imputing missing data to assess the associations of several molecular features with PCCRCs compared to DCRCs. Multiple imputation of missing data allows for missingness depending on observed variables (missing at random; MAR), uses all available data (no list-wise deletion), and reduces the risk of bias from features that coincide with lesser quantity of tissue available for molecular analysis.

The missingness of molecular features was imputed using the other molecular features and patient characteristics (with correlation >= 0.01) as predictors. The patient characteristics consisted of gender, patient age, tumour location, early stage, mucous CRC, morphology (polypoid/non-polypoid), size, presence of diverticulosis and whether the CRC was a PCCRC or not. The MICE package version 2.46.0 was used to impute missing data, using 30 sets with 20 iterations.^29^ Convergence was checked by inspecting the trace lines.

Unsupervised hierarchical clustering was performed on a binary distribution of molecular features. All molecular features which appeared to be different between groups in univariate analysis of both all PCCRC and biological PCCRC analysis, were included. Additionally, all mutations with an observed prevalence of minimal 9% were included.

The Ward.D algorithm of the *hclust()* function in R statistics was used for clustering.^30^ This is a distance algorithm finding compact and spherical clusters. It is similar to a complete algorithm that takes the lowest sum of squared distances of the average in a cluster.^31,32^ It is a commonly used algorithm when there is no specific hypothesis about linkage between the observations in advance. This was the case with these data.

Heatmaps were plotted using Gplots.^33^ The heatmaps show patterns which are in line with the known subtypes as published previously.^34^ So, the use of the Ward algorithm seems legitimate with these data. The clusters were cut based on the same previously published subtypes of CRC and corresponding molecular features.

Based on the dendrogram, the number of primary branches was determined. Differences in the proportion of PCCRCs and genetic alterations between branches were tested using the Chi-square test. All statistical analyses were performed with R statistics version 3.4.0.^30^

## Results

During a 10-years period, 5701 patients were diagnosed with CRC within the South Limburg region. Of these patients, 594 were excluded because of hereditary CRC, IBD or a previous history of CRC (Figure 1). The remaining 5107 patients had a total of 5303 CRCs, of which 151 were PCCRCs according to the WEO classification. From the remaining DCRCs, 143 controls were randomly selected. High quality DNA was available in 122 of 151 PCCRCs and 98 of 143 DCRCs. CIMP status was available for all samples. Good quality DNA copy number profiles were obtained for 105/122 PCCRCs and 88/98 DCRCs. In some cases, DNA was insufficient for analysis of MSI status and mutation data (Figure 1).

**Figure 1:**
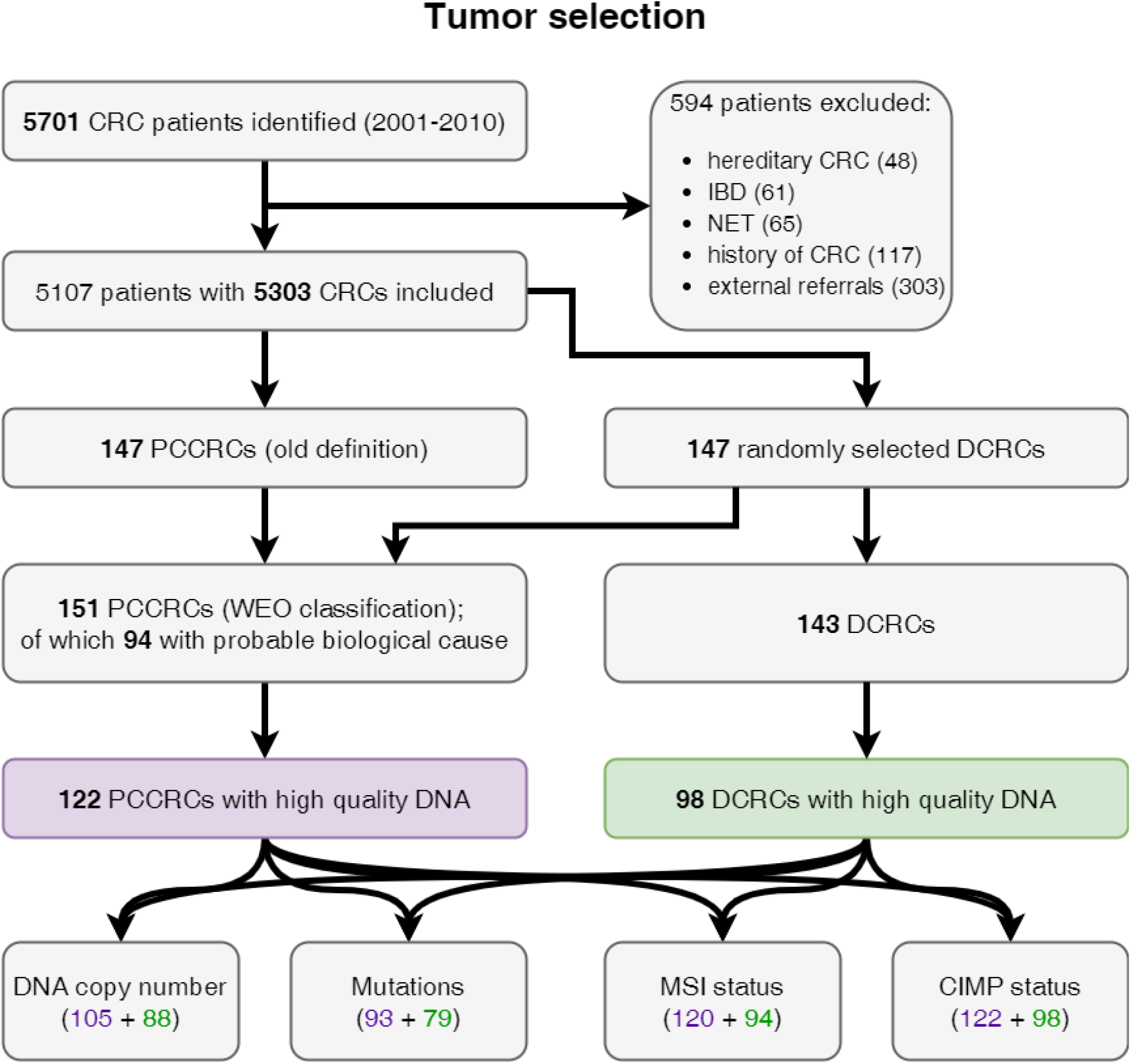
Flow-chart of the process of selection of CRC cases for molecular analysis. PCCRCs, post-colonoscopy CRCs; DCRCs, detected CRCs.

Of the 122 PCCRCs used in molecular analysis, 94 had a probable biological cause (75 cases of possible missed lesions with prior adequate examination, and 19 cases of likely new CRC) and 28 had a probable procedural cause (21 cases of possible missed lesions with prior inadequate examination, 6 cases of likely prior incomplete resection, and 1 case of previous detected lesion without resection).

### Clinical characteristics

Clinical characteristics of the two groups of CRC patients are shown in Table 1. Baseline characteristics of the PCCRCs were significantly different in several aspects from DCRCs with respect to proximal location, flat appearance, and smaller size (Table 1). PCCRCs were significantly more often stage T1 carcinoma and poorly differentiated compared to DCRCs.

**Table 1:**
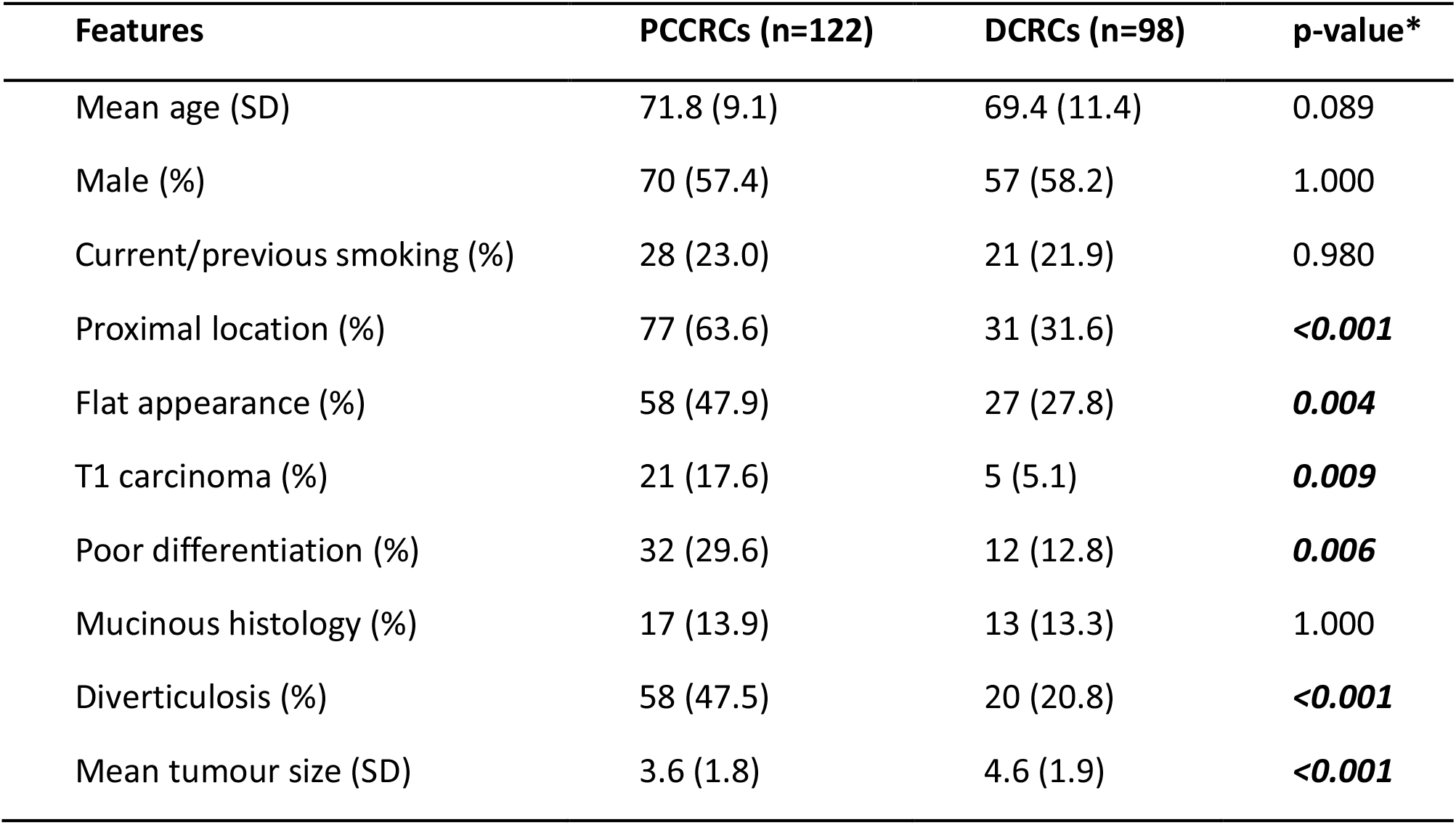
Baseline characteristics of PCCRCs versus DCRCs. * P-value < 0.05 considered significant.

| Features | PCCRCs (n=122) | DCRCs (n=98) | p-value* |
| --- | --- | --- | --- |
| Mean age (SD) | 71.8 (9.1) | 69.4 (11.4) | 0.089 |
| Male (%) | 70 (57.4) | 57 (58.2) | 1.000 |
| Current/previous smoking (%) | 28 (23.0) | 21 (21.9) | 0.980 |
| Proximal location (%) | 77 (63.6) | 31 (31.6) | **<0.001** |
| Flat appearance (%) | 58 (47.9) | 27 (27.8) | **0.004** |
| T1 carcinoma (%) | 21 (17.6) | 5 (5.1) | **0.009** |
| Poor differentiation (%) | 32 (29.6) | 12 (12.8) | **0.006** |
| Mucinous histology (%) | 17 (13.9) | 13 (13.3) | 1.000 |
| Diverticulosis (%) | 58 (47.5) | 20 (20.8) | **<0.001** |
| Mean tumour size (SD) | 3.6 (1.8) | 4.6 (1.9) | **<0.001** |

### Molecular features of all PCCRCs versus DCRCs

DNA copy number analysis was complete in 105 PCCRC cases (86.1%) and 88 DCRC cases (89.8%) (Figure 1). Overall, PCCRCs and DCRCs had comparable patterns of chromosomal alterations (gains and losses), with high prevalence of gains of 7p, 7q, 8q, 13q, 20q and losses of 8p, 17p, 18p and 18q (Supplementary Figure 1). However, PCCRCs showed less frequently loss of 18q in comparison to DCRCs in univariate CGH analysis (FDR <0.2).

Mutation data were complete in 93 PCCRC cases (76.2%) and in 79 DCRC cases (80.6%) (Figure 1). A panel of 48 cancer-related genes was tested (Supplementary Figure 2), of which none of the genes was significantly different between PCCRC and DCRC in univariate testing. The results focus on the nine altered genes with prevalence of at least 9% in all CRCs analysed, namely *APC, BRAF, FBXW7, KIT, KRAS, PIK3CA, PTEN, SMAD4* and *TP53*, see Table 2. Gene mutation frequencies were comparable between PCCRCs and DCRCs.

**Table 2:** Molecular characteristics of PCCRCs versus DCRCs. * P-value < 0.05 considered significant; ** FDR<0.2 considered significant.

| Features | PCCRCs (n=122) | dCRCs (n=98) | p-value* | FDR** |
| --- | --- | --- | --- | --- |
| APC gene mutation | 37/93 (39.8) | 39/79 (49.4) | 0.268 |  |
| BRAF gene mutation | 17/93 (18.3) | 8/79 (10.1) | 0.195 |  |
| FBXW7 gene mutation | 8/93 (8.6) | 9/79 (11.4) | 0.723 |  |
| KIT gene mutation | 19/93 (20.4) | 18/79 (22.8) | 0.851 |  |
| KRAS gene mutation | 32/93 (34.4) | 24/79 (30.4) | 0.690 |  |
| PIK3CA gene mutation | 16/93 (17.2) | 13/79 (16.5) | 1.000 |  |
| PTEN gene mutation | 11/93 (11.8) | 6/79 (7.6) | 0.502 |  |
| SMAD4 gene mutation | 8/93 (8.6) | 9/79 (11.4) | 0.723 |  |
| TP53 gene mutation | 36/93 (38.7) | 38/79 (48.1) | 0.278 |  |
| Gain of chromosome 13q | 52/105 (49.5) | 60/88 (68.2) | 0.014 | <0.303 |
| Loss of chromosome 17p | 45/105 (42.9) | 42/88 (47.7) | 0.595 | <0.986 |
| Loss of chromosome 18q | 49/105 (46.7) | 64/88 (72.7) | <0.001 | <0.107 |
| MSI | 26/120 (21.7) | 9/94 (9.6) | 0.029 | - |
| CIMP high profile | 61/122 (50.0) | 32/98 (32.7) | 0.014 | - |

**Figure 2:**
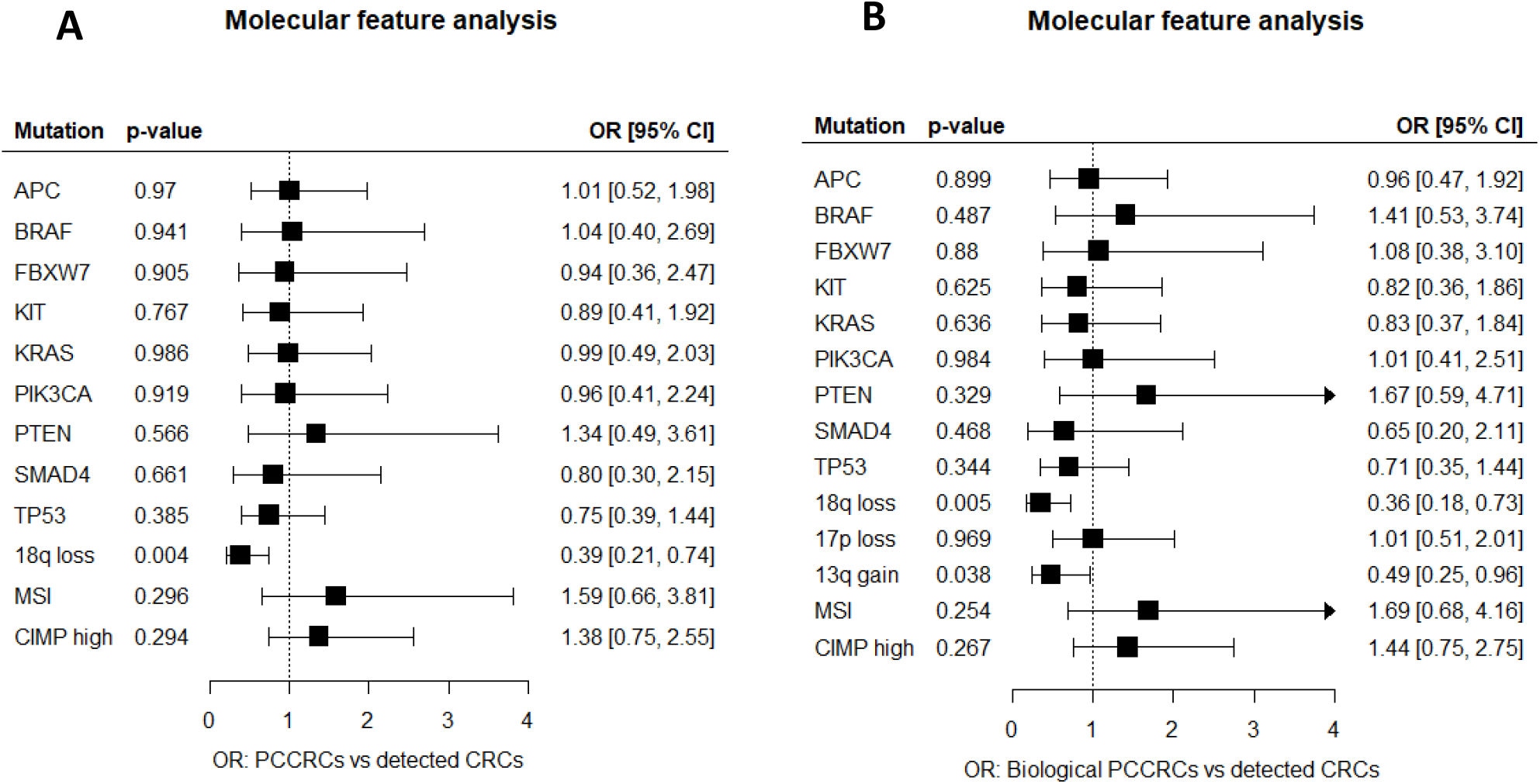
Comparison of molecular features between post-colonoscopy colorectal cancers (PCCRCs) and detected colorectal cancers (DCRCs). Forest plots showing the associations of several molecular features of: A) PCCRCs compared to DCRCs, and B) PCCRCs with plausible biological aetiology compared to DCRCs, after multiple imputation of missing values and correction for age, gender and tumour location. OR, Odds Ratio; CI, confidence interval.

#### MSI and high CIMP status were significantly more common in PCCRCs vs DCRCs (Table 2)

To correct for partially missing molecular data, multiple imputation (MI) was used (Supplementary Table 1). A logistic regression model analysis was performed after MI, corrected for gender, age at diagnosis and tumour location, and the obtained pooled estimation ORs and 95% CIs were similar to those obtained from complete case analysis (list-wise deletion of cases with missing values). After applying correction for age, gender, and tumour location, only loss of 18q chromosome remained significantly less common in the PCCRCs (OR 0.4, 95%CI: 0.2-0.7; Figure 2A).

### Molecular features of biological PCCRCs only versus DCRCs

When comparing the 94 PCCRCs caused by biological factors to the DCRCs, next to loss of 18q chromosome, gain of the 13q chromosome and loss of a small segment of the 17p chromosome (1 MB size), were also significantly less often present in PCCRCs compared to DCRCs in univariate analysis (Table 3). MSI and high CIMP remained significantly more prevalent in PCCRCs. In the 48 gene panel, significantly more BRAF was present in PCCRCs compared to DCRCs (p=0.049 and p=0.031 respectively). After correction for age, gender and tumour location in a logistic regression model, of the above mentioned differences, only 18q loss and 13q gain remained significantly different. All other molecular characteristics between PCCRCs and DCRCs were comparable (Figure 2B).

**Table 3:**
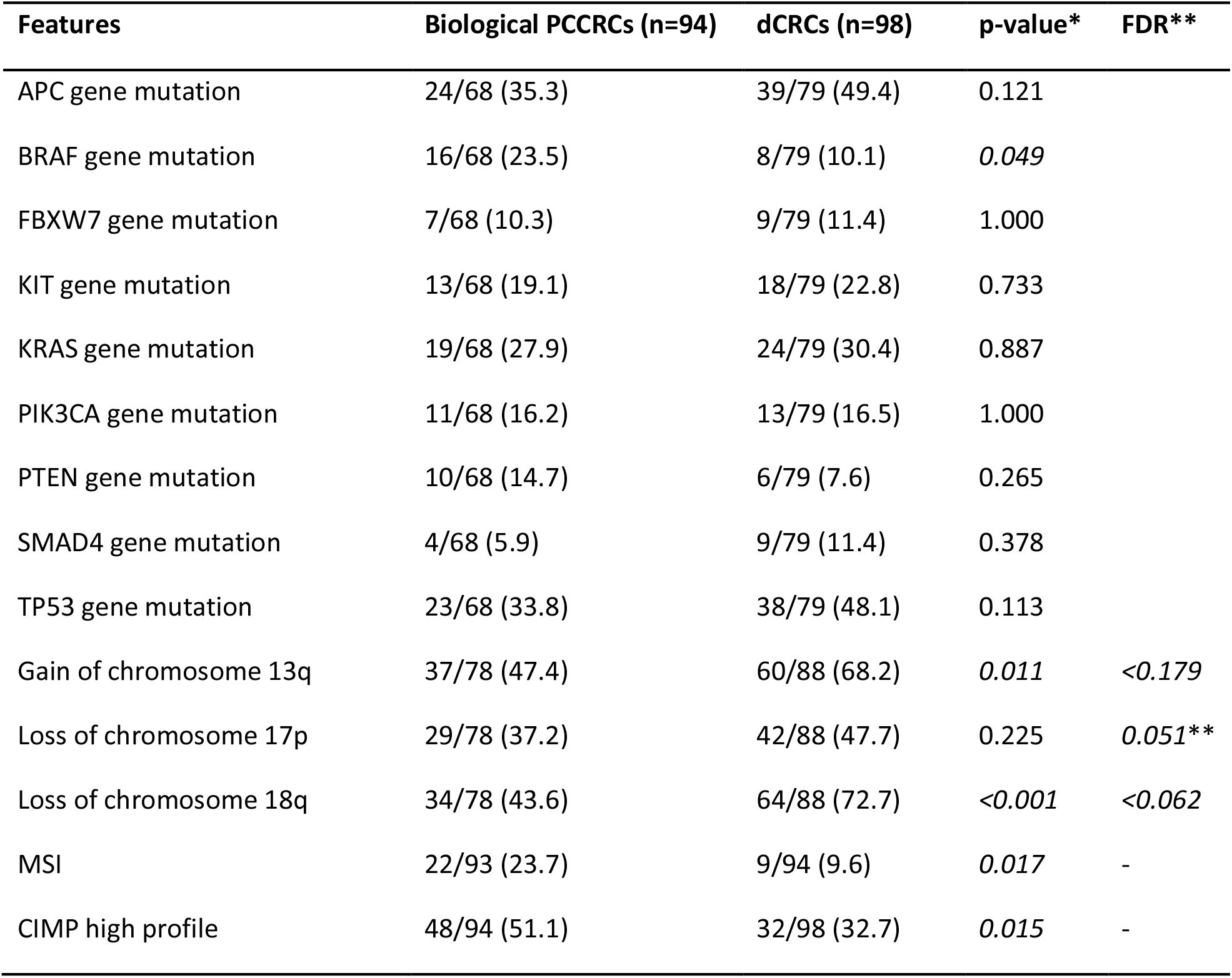
Molecular characteristics of PCCRCs with likely biological cause versus DCRCs. * P-value < 0.05 considered significant; ** FDR<0.2 considered significant; ^#^ Fisher exact test; ^(1)^1 MB size segment (19.1-20.1 MBp)

### Clustering analysis considering all molecular features

Interaction between the molecular features was not tested in the logistic regression models because of limited power. Hierarchical clustering was used to show whether certain molecular features correlated. The prevalence of each CRC type within the identified clusters was tested afterwards.

In selecting variables for the clustering analysis, loss of 18q, loss of 17p and gain of 13q were included since these had at least one significant segment difference in univariate analysis. Additionally, MSI and CIMP were included because of univariate significance. Finally, we added the 9 gene mutations with a prevalence of ≥9% (APC, BRAF, FBXW7, PIK3CA, SMAD4, TP53, KRAS, PTEN and KIT mutations). With hierarchal clustering, 3 major branches of CRCs were detected (Figure 3a). Within the branches, the prevalence of PCCRCs was significantly different: 62.0%, 67.9% and 46.5% (p=0.018) for branch 1, 2 and 3, respectively. Biological PCCRCs were also observed more frequently in clusters 1 and 2, compared to 3 (48.0% and 53.6% compared to 35.1%; p value = 0.010) (Figure 3a, Supplementary Table 2a). Branch 1 was characterized by the presence of high CIMP (100%), with only 1 case of MSI (2.0%) and relatively frequent *BRAF* mutations (25.6%) (Figure 3b, Supplementary Table 2b). Branch 2 had frequently MSI (56.4%) and a high frequency of *BRAF* mutations (31.9%), and very often high CIMP (60.7%). Finally, branch 3 had hardly any MSI (2.7%), a few cases of high CIMP (7.9%) and no *BRAF* mutations (0.0%), and it was enriched for DNA copy number changes (p<0.001). Gain of chromosome 13q (77.9%), loss of chromosome 17p (68.4%) and loss of chromosome 18q (82.1%) were most common in branch 3, but also in branch 1 (63.8%, 44.7% and 70.2%, respectively) (Figure 3). The combination of high CIMP and MSI appeared to be most associated with PCCRCs (Figure 3b). PCCRCs had significantly more often both CIMP and MSI than detected CRCs (15.7% vs 4.1%, p=0.010). The prevalence of non-polypoid CRCs was not significantly different between branches (p=0.073). Proximal location clearly differed among the clusters, being more prevalent in branches 1 and 2 compared to branch 3 (p<0.001).

**Figure 3:**
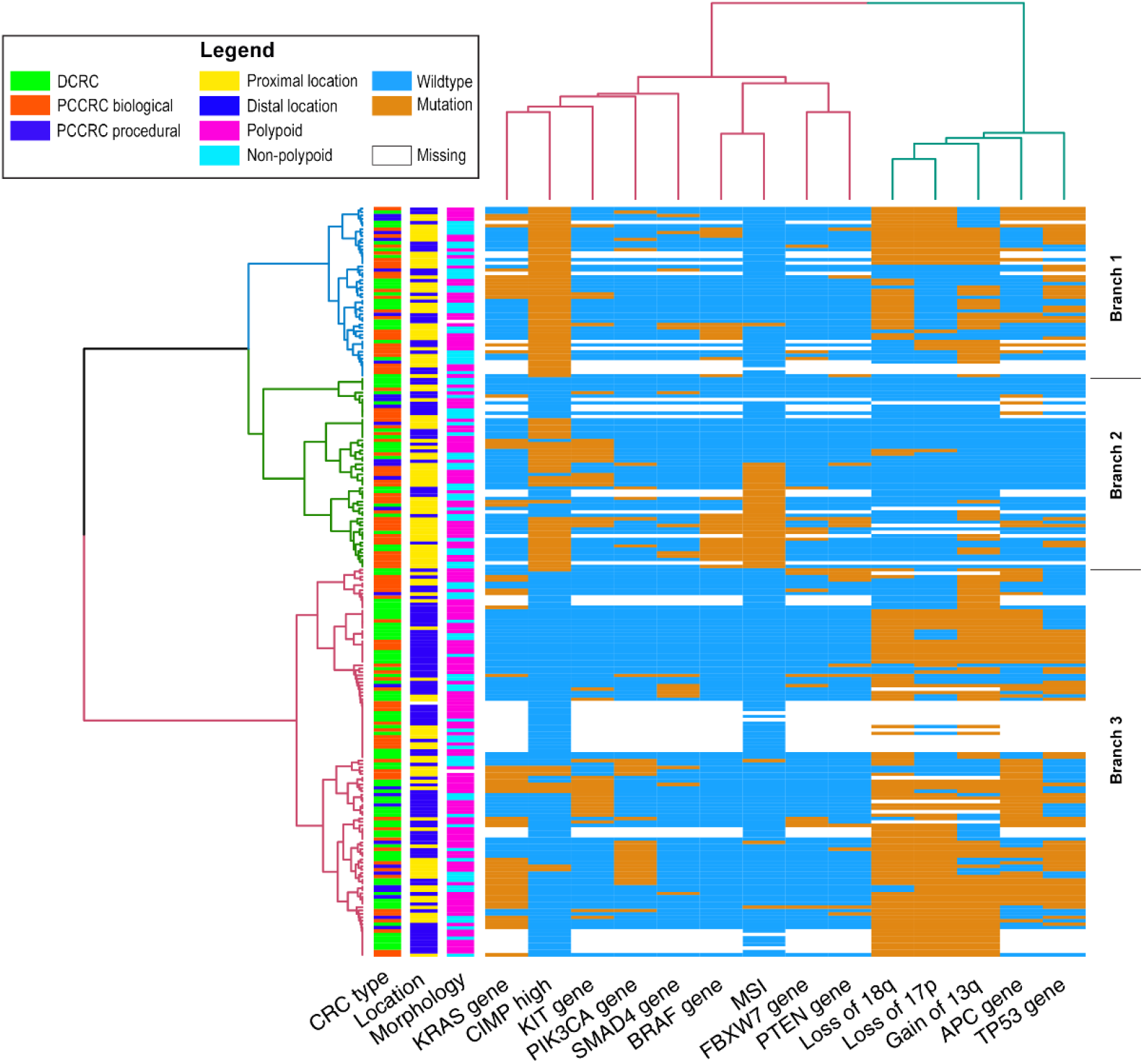
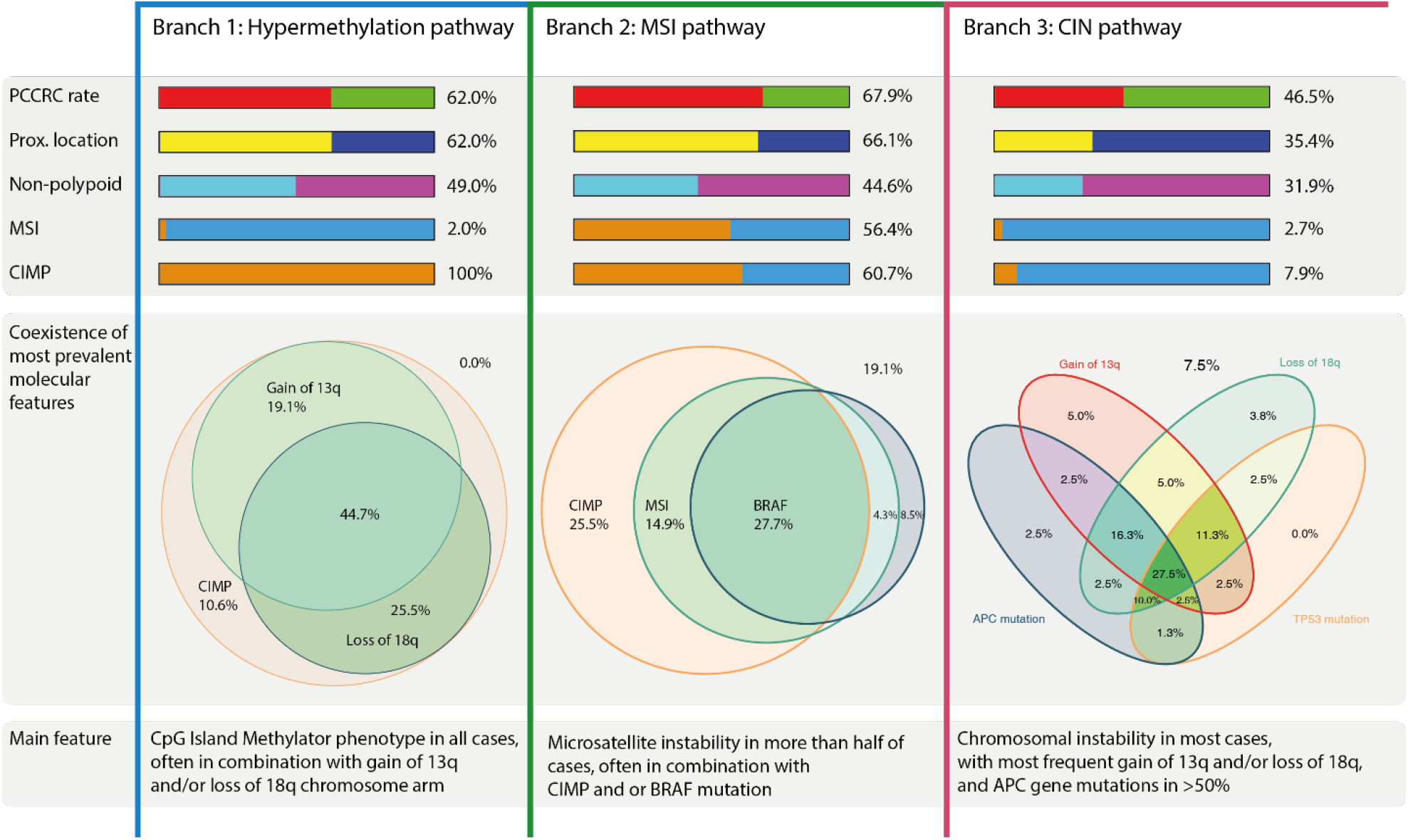
Unsupervised hierarchal cluster analysis based on the molecular features of all CRCs analysed. In the cluster analysis results of the nine genes most commonly mutated in this study (APC, TP53, KRAS, KIT, PIK3CA, BRAF, FBXW7, SMAD4 and PTEN), the significant chromosomal alterations (loss of 17p, loss of 18q and gain of 13q), CIMP status and MSI, were included. A) Heatmap displaying the distribution of all clinical and molecular features of all CRCs analysed in this study. Orange indicates presence while blue indicates absence of these features. The first three columns represent CRC type (Biological PCCRCs: red, Procedural PCCRCs: blue, DCRCs: green), CRC location (proximal: yellow, distal: dark blue) and CRC morphology (polypoid: purple, non-polypoid: light blue). After clustering of the CRCs, three large branches can be detected (blue, green, and red). B) Overview of hallmark features of each branch from clustering analysis. Red: PCCRC, green: DCRC, yellow: proximal location, dark blue: distal location, light blue: non-polypoid, purple: polypoid, orange: MSI / high CIMP present, blue: MSI / high CIMP absent.

## Discussion

We analysed molecular features of PCCRCs and of DCRCs in a nested case-control series derived from a well-defined population-based cohort, to test the hypothesis whether PCCRCs have a molecular profile that is different from DCRCs. In addition to MSI and CIMP status, which have been frequently analysed in previous studies, we used extensive profiling including low-coverage whole genome sequencing to determine DNA copy number alterations as well as targeted next-generation sequencing, targeting a panel of 48 cancer-related genes, to test our hypothesis.

PCCRCs were significantly more often located in the proximal colon, had more often a flat appearance, were more often smaller in size and more often contained early CRC. Molecular analysis showed that PCCRCs were more frequently MSI and CIMP-high and showed less frequently losses of chromosome arm 18q, when compared to DCRCs. However, after correction for age, gender and tumour location, only loss of the 18q chromosome arm remained significant as PCCRCs are strongly correlated with proximal location. Considering only PCCRCs with a possible biological cause in the comparison to DCRCs, we observed that PCCRCs were more MSI, with high CIMP, had more frequently BRAF mutations, had less frequently losses of 18q and 17p as well as less gain of 13q. However, again, after correction for age, gender and tumour location, only loss of 18q and gain of 13q remained significant. Of interest, no significant differences in any molecular features were found between procedural PCCRCs and DCRCs, as expected (data not shown).

Previous studies, comparing PCCRCs with DCRCs found higher rates of MSI and CIMP among PCCRCs,^14,15^ although in some studies the prevalence of MSI among PCCRCs compared to DCRCs was only moderately increased.^18,35^ This is in line with our findings, where we observed a significant difference in MSI and CIMP status that disappeared when we corrected for tumour location. A study looking at PCCRCs diagnosed within 5 years after negative colonoscopy, showed no difference between *BRAF, KRAS* and *PIK3CA* gene mutations.^16^ These gene mutations were also in our study not statistically different between PCCRCs and DCRCs.

For the first time, we show that certain DNA copy number alterations are less frequent in PCCRC compared to DCRC, and that is even more clear when considering only biological PCCRCs in the comparison. However, we do observe a similar whole genome DNA copy number pattern in PCCRCs compared to DCRCs, implying that chromosomal instability (CIN) also plays a role in PCCRCs. This is also in agreement with a previous study showing that interval CRCs presented a CIN phenotype, similar to non-interval CRCs, although this analysis was based on FISH data comprising only chromosomes 8, 11 and 17.^36^

Multiple pathways for the development of CRC have been proposed. Chromosomal instability (CIN), microsatellite instability (MSI) and hypermethylation are considered the three main pathways, although these pathways are not fully independent of each other.^37-40^ Our cluster analysis, considering all significant variables (univariate analysis) and gene mutations occurring in at least 9% of the cases, showed three major clusters with marked similarity to these pathways. Cluster 1 was characterized by CIMP and DNA copy number changes, with almost no MSI, reflecting the *hypermethylation pathway*. Cluster 2 was characterized by MSI with frequent CIMP and *BRAF* gene mutations and reflects the *MSI pathway*. Cluster 3 was characterized by absence of high CIMP and MSI and by a high frequency of DNA copy number changes and could be considered as the *CIN pathway*. While cluster 3 contained most CRCs, in cluster 2 the percentage of PCCRCs was highest (67.9%).

*BRAF* mutations were not independently associated with PCCRCs, but were shown to have a strong association with MSI in another study.^41^ Our clustering analysis indeed shows one cluster with frequent MSI and another with frequent CIMP high cases independent of MSI status. Our data confirm the strong association between PCCRCs and MSI, *BRAF* gene and CIMP phenotypes, but a specific PCCRC pathway was not found. The association between location and presence of MSI, CIMP and BRAF mutations has been studied before and is hypothesized to explain the lower efficacy of proximal colonoscopy.^42^ The majority of the PCCRCs included in this study were considered to result from missed lesions with previously performed adequate examination (75/122 PCCRCs).

PCCRCs have been associated with non-polypoid and sessile serrated precursor lesions, so their molecular profiles may be more similar. Sessile serrated lesions (SSL) are considered potential contributors to PCCRCs because of their flat or sessile appearance with pale colour, high prevalence in the proximal colon and their subtle lesion borders that make radical resection more difficult.^43,44^ CIMP and *BRAF* mutations and to a lesser degree MSI are associated with the pathogenesis of SSLs.^45,46^ Our data reveal that in the PCCRC group CIMP high profile, BRAF and CIMP are more prevalent compared to the DCRC group, supporting an association with SSLs. Non-polypoid colorectal neoplasms (CRNs) in general have a molecular profile that is different from polypoid CRNs.^47^ DNA copy number losses of chromosome 17p and 18q were observed less frequently in non-polypoid vs polypoid CRN.^24^ Mutations in *KRAS* and *APC* were less common while BRAF mutations were more common in non-polypoid than in polypoid CRNs.^47,48^ No differences in MSI status were observed^48,49^ and evidence on differences in CIMP status is lacking. Several of these molecular features (KRAS, APC, BRAF mutations, DNA copy number changes) correspond with features that we identified in PCCRCs. Therefore, based on similarities in molecular profiles, our data support the hypothesis that both SSLs and non-polypoid CRNs may contribute to development of PCCRC.

In order to reduce the occurrence of PCCRC, detection and determination of non-polypoid CRN and SSLs should be improved. Detailed training of endoscopists in recognizing non-polypoid CRN and SSLs is important and has been proven to be successful.^50^ The introduction of benchmarks and of training has resulted in increased adenoma detection rates^51-53^ and has decreased the risk on PCCRCs.^54,55^ In the near future, technical advances like artificial intelligence, may help to improve detection, determination and adequate endoscopic resection of subtle colorectal neoplasms and thereby help to further reduce the percentage of PCCRCs.^56^

Some limitations of our study should be acknowledged. First, not all CRC samples harboured high quality DNA, leading to missing data in the molecular analyses. To overcome this, multiple imputation analysis was used for the results, showing no differences with the complete case analysis. Second, clinical data were collected retrospectively, based on patient files and national and regional registries. Reliability of the results depends on completeness of these registries and of the patient records. To limit bias, cross-reference checks have been performed.^5^ Migration in and out of the region could lead to undetected PCCRCs. However, the migration rate in the South Limburg region is very low and all three hospitals in the region were included. Third, while patients with known Lynch syndrome were excluded, some yet undiagnosed cases may be present in the included cases. However, the majority of cases with MSI are likely sporadic (simultaneous BRAF mutation/CIMP profile) since only one case of MLH1 mutation occurred. Lastly, we used a 1 on 1 ratio in selecting PCCRCs and DCRCs instead of a larger control group. This could have reduced the power of the study.

Strengths of our study are that to our knowledge it is the first study, a) integrating whole genome DNA copy number sequencing with CRC mutation analysis and MSI and CIMP status, b) with unsupervised hierarchical clustering analysis to form unbiased groups, and c) with cases and controls selected for this study derived from a well characterized population-based cohort with detailed information on probable aetiology.

## Conclusion

Compared to detected CRCs, PCCRCs are significantly more often proximally located, non-polypoid appearing, early stage and poorly differentiated. PCCRCs showed molecular characteristics common to the canonical CIN, MSI and hypermethylation pathways. After correction for gender, age at diagnosis and tumour location, PCCRCs compared to detected CRCs harboured less often loss of 18q chromosome. Although no PCCRC specific pathway could be defined, pathways associated with sessile serrated and non-polypoid CRNs were more common. In combination with the clinical features observed in PCCRCs, these findings support the hypothesis that SSLs and non-polypoid CRNs are contributors to the development of these cancers. In order to further reduce the occurrence of PCCRC, the focus should be directed at improving the detection, determination and endoscopic removal of these non-polypoid CRN and SSLs.

## Data Availability

The sequencing data used in the present study is available in the European Genomes and phenomes Archive (ega-archive.org), with provisory number EGAS00001004686.

## Abbreviations

95%CI: 95% confidence interval
CIMP: CpG Island Methylator Phenotype
CRC: Colorectal cancer
CIN: Chromosomal Instability
CRN: Colorectal neoplasm
DCRC: Detected CRC
FFPE: Formalin-fixed, paraffin-embedded
MSI: Microsatellite instability
OR: Odds ratio
PCCRC: Post-colonoscopy colorectal cancer
SSL: Sessile serrated lesion
WGS: Whole genome sequencing

## Additional information

## Acknowledgements

We thank the technicians of the pathology laboratory at the Netherlands Cancer Institute in Amsterdam for their help with the DNA isolations. We thank the Tumour Genome Analysis Core at Amsterdam UMC for low-coverage WGS and cancer-related gene panel targeted sequencing. We thank the Genomics Core Facility at the Netherlands Cancer Institute for cancer-related gene panel targeted sequencing. We thank the technicians of the pathology laboratory at the Maastricht University Medical Centre for their help with the CIMP analysis. We thank dr. Silvia Sanduleanu for her contribution to the origin of this study.

We acknowledge that a part of this work has been presented at the UEGW 2016 in Vienna (Bogie RMM, Clercq CMC le, Voorham QJM, Cordes M, Sie D, Broek E vd et al. OP212 ASSOCIATION OF CHROMOSOMAL INSTABILITY AND MICROSATELLITE INSTABILITY PATHWAYS WITH POSTCOLONOSCOPY COLORECTAL CANCER IN A RETROSPECTIVE COHORT STUDY. United European Gastroenterology Journal 2016; 4(5_suppl): A1-A156 [Oral presentation 212]). This work was performed within the frame of the COST Action [CA17118], supported by COST (European Cooperation in Science and Technology).

## Author’s contributions

RB, CC, ME, GM and BC conceived the study concept and design; RB, CC, QV, MC, DS, CR, EB, SV, NG, RR, PS, ES, BY and BC contributed to the acquisition of data; RB, CC, QV, MC, DS, CR, EB, SV, NG, RV, BW, ME, BY, GM, AM and BC participated in analysis and interpretation of data; RB, CC, QV, CR, BW, AM and BC drafted the manuscript; all authors did a critical revision of the manuscript for important intellectual content; RB, CR, RV and BW performed the statistical analysis; AM and BC had the study supervision. AM is the guarantor of the article. All authors approved the final version.

## Ethics approval and consent to participate

The study was approved by the Medical Ethical Committee of the Maastricht University Medical Centre, which waived the need for informed consent. The study is registered as study NTR3093 in the Dutch trial register (www.trialregister.nl). The study was performed in accordance with the Declaration of Helsinki.

## Consent for publication

No individual data was presented in the current study.

