## Supplementary data for "Molecular pathways in post-colonoscopy versus detected colorectal cancers: results from a nested case-control study"

**PCCRCs**

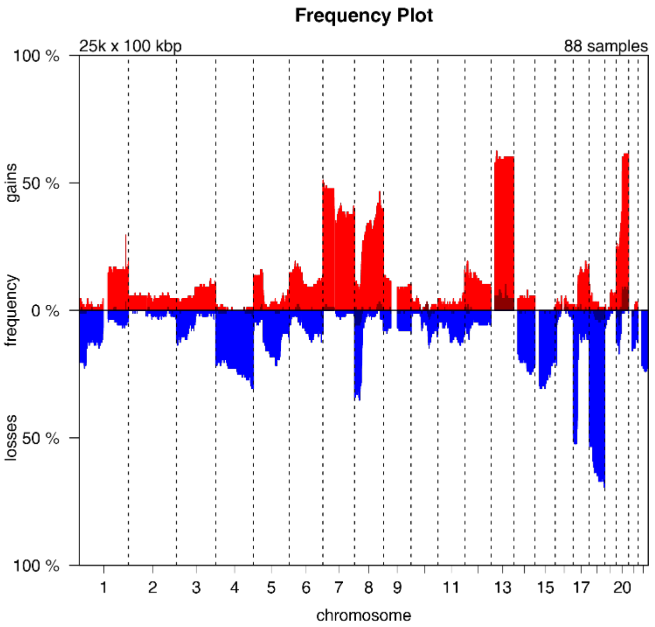


**DCRCs**

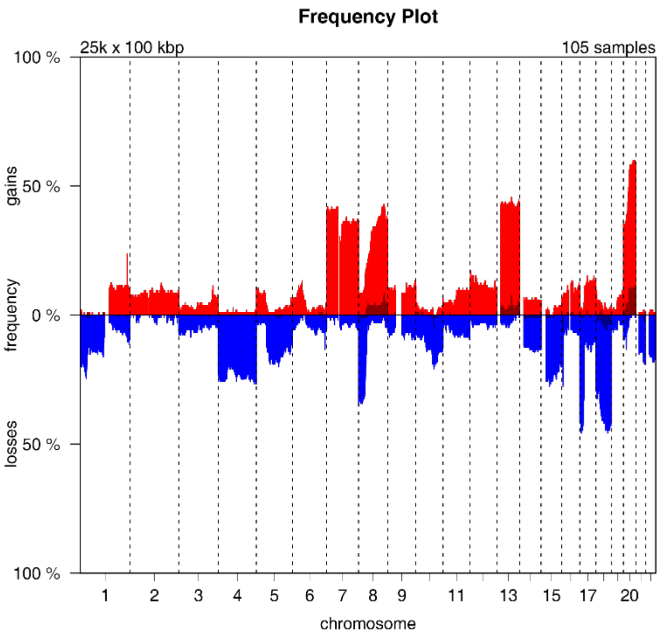


Supplementary Figure 1: PCCRCs and DCRCs frequency plots of DNA copy number gains and losses throughout the whole genome. Red: gains, blue: losses.

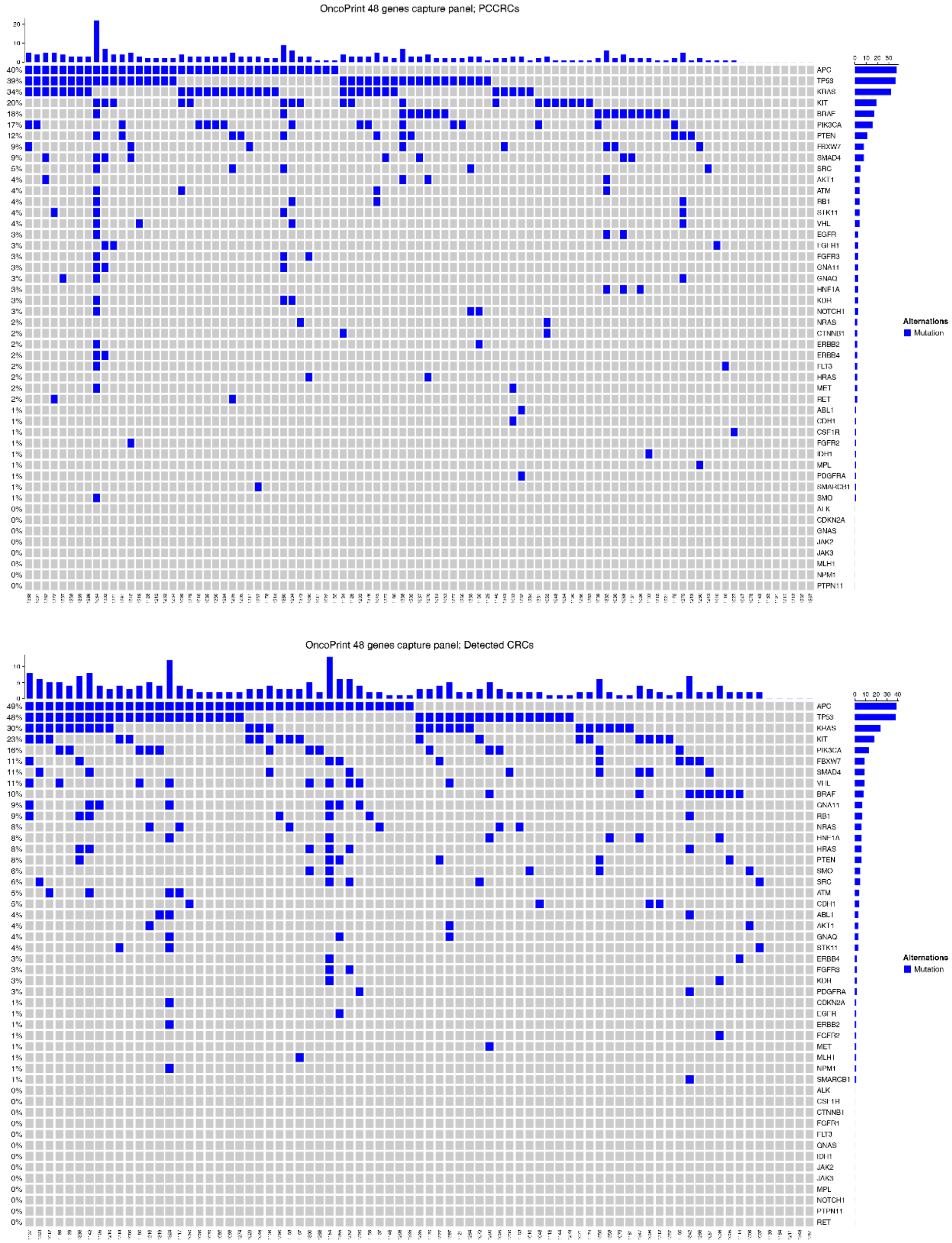


Supplementary Figure 2: Mutation frequencies detected in PCCRCs and DCRCs. Oncoprint matrix showing the 48 tested genes (rows) in CRC cases (columns). Blue coloured block: mutations detected; grey coloured block: no mutation detected. Top panel: results of PCCRCs; lower panel: results DCRCs.

Supplementary Table 1: Comparison of baseline characteristics between CRC cases with and without missing data. CCA: complete case analysis; *Size is missing in 4 cases.

| Characteristic | CCA (n=162) | Imputed cases (n=58) | p-value |
|---|---|---|---|
| PCCRC (%) | 89 (54.9) | 33 (56.9) | 0.918 |
| Mean age in years (SD) | 71.0 (10.4) | 70.0 (9.7) | 0.499 |
| Mean CRC size in cm (SD) | 4.07 (1.88) | 4.05 (1.99) | 0.959 |
| Gender (% male) | 93 (57.4) | 34 (58.6) | 0.996 |
| Location CRC (% proximal) | 85 (52.5) | 23 (40.4) | 0.156 |
| Morphology (% flat) | 66 (41.3) | 19 (32.8) | 0.328 |
| TNM stage (% T1) | 18 (11.1) | 8 (14.5) | 0.662 |
| CRC differentiation (% good/intermediate) | 119 (77.8) | 39 (79.6) | 0.945 |
| Mucinosity (% >50% present) | 19 (11.7) | 11 (19.0) | 0.248 |

Supplementary Table 2a+b: Comparison of branches of the hierarchical clustering analysis. *Chi-square test used to compare whether PCCRC rates were different between branches. ** Chi-square test used to compare whether the subset biological PCCRC was different among branches.

A)

| Branch | PCCRC (55.5%) | Biological PCCRC (42.7%) | Procedural PCCRC (12.7%) | Proximal location | Nonpolypoid |
|---|---|---|---|---|---|
| 1 (n=50) | 31 (62.0%) | 24 (48.0%) | 7 (14.0%) | 31 (62.0%) | 24 (49.0%) |
| 2 (n=56) | 38 (67.9%) | 30 (53.6%) | 8 (14.3%) | 37 (66.1%) | 25 (44.6%) |
| 3 (n=114) | 53 (46.5%) | 40 (35.1%) | 13 (14.3%) | 40 (35.4%) | 36 (31.9%) |
| | P=0.018* | P=0.084** | | P<0.001* | P=0.073* |

B)

| Branch | MSI | CIMP high | BRAF | KRAS | 13q gain | 17p loss | 18q loss |
|---|---|---|---|---|---|---|---|
| 1 (n=50) | 1 (2.0%) | 50 (100%) | 10 (25.6%) | 13 (33.3%) | 30 (63.8%) | 21 (44.7%) | 33 (70.2%) |
| 2 (n=56) | 31 (56.4%) | 34 (60.7%) | 15 (31.9%) | 6 (12.8%) | 8 (15.7%) | 1 (2.0%) | 2 (3.9%) |
| 3 (n=114) | 3 (2.7%) | 9 (7.9%) | 0 (0.0%) | 37 (43.0%) | 74 (77.9%) | 65 (68.4%) | 78 (82.1%) |
